# DraculR: A web based application for *in silico* haemolysis detection in high throughput small RNA sequencing data

**DOI:** 10.1101/2022.03.27.22273019

**Authors:** Melanie D. Smith, Shalem Y. Leemaqz, Tanja Jankovic-Karasoulos, Dylan McCullough, Dale McAninch, James Breen, Claire T. Roberts, Katherine A. Pillman

## Abstract

**Motivation:** The search for novel microRNA (miRNA) biomarkers in plasma is hampered by haemolysis, the lysis and subsequent release of red blood cell (RBC) contents, including miRNAs, into surrounding fluid. The biomarker potential of miRNAs comes in part from their multi-compartment origin, and the long-lived nature of miRNA transcripts in plasma, giving researchers a functional window for tissues that are otherwise difficult or disadvantageous to sample. The inclusion of RBC derived miRNA transcripts in downstream analysis introduces a source of error that is difficult to identify *post hoc* and may lead to spurious results. Where access to a physical specimen is not possible, our tool will provide an *in silico* approach to haemolysis prediction.

**Results:** We present DraculR, an interactive Shiny/R application that enables a user to upload microRNA expression data from short read sequencing of human plasma as a raw read counts table and interactively calculate a metric that indicates the degree of haemolysis contamination.

**Availability and implementation:** DraculR and its tutorial are freely available from (https://mxhp75.shinyapps.io/shinyVamp/). Code is available from (https://github.com/mxhp75/shinyVamp.git).

## Introduction

Circulating miRNAs have long been identified in human plasma and, given their stability in this medium, have strong potential as biomarkers. Whilst there are multiple techniques for quantifying the abundance of miRNAs in plasma, high throughput sequencing (HTS) detects both known and novel (ie. putative) miRNAs with single base resolution. The fine resolution provided by HTS allows distinction between variants differing by a single nucleotide, as well as isomiRs of differing length (1) with many researchers now leveraging this technology (2–11). The biomarker potential of plasma miRNAs is in part because plasma derived transcripts commonly originate from varied endogenous compartments. Accurate profiling of plasma miRNAs is impeded when transcripts are derived from another blood source such as RBCs. When haemolysis occurs due to shearing of RBCs during blood sampling, miRNAs are released into the volume of blood drawn (12–16). The presence of the RBC-associated miRNAs alters the plasma expression profile affecting the global normalisation of sequence counts (1).

The increase in relative abundance of RBC-associated miRNAs, and the aberrant normalisation of libraries, have potential to impact the profile analysis of miRNAs (12,13,15) yet assessment of haemolysis is rarely reported. Data quality checks prior to analysis of HTS data should include an assessment of haemolysis in the plasma sample from which the sequencing library was produced. There are currently two gold standard approaches: 1. Delta quantification cycle (ΔCq), where expression levels of a known blood cell associated miRNA (miR-451) and a control miRNA (miR-23a) are determined based on the difference between the two raw Cq values; and 2. Spectrophotometry, based on absorbance maximum of free haemoglobin measured at 414 nm (12,17–19). However, both gold standard methods rely on access to the original plasma sample. In this work we present DraculR, a data driven approach for the assessment of haemolysis confounding *in silico*. The DraculR tool enables the user to upload self-generated or publicly available high throughput miRNA sequencing data for assessment and returns both visual and tabular recommendations for downstream analysis of read count data.

## Materials and methods

DraculR is an interactive, Shiny/R web-based tool, for the *in silico* assessment of haemolysis contributions to small RNA sequencing libraries prepared from human plasma. DraculR utilises the Haemolysis metric (20), a measure analogous to the ΔCq (miR-23a-miR-451) method, which determines the difference between the abundance of two miRNAs, one known to vary, and one known to be invariant in the presence of haemolysis. The Haemolysis metric is calculated as the sample specific difference in geometric means of the normalised gene expression values between two sets of microRNAs: 1) 20 miRNAs identified as indicative of haemolysis (‘signature set’), and 2) all other microRNAs (‘background’). In this case, the geometric mean of the reduced signature set will be calculated, as defined in (1).

Let

*Z*_*x*_ be the miRNA Reduced signature set (log_2_ CPM counts)

and *Z*_*y*_ be the Background miRNA set (log_2_ CPM counts)

where *x* = 1,2,3, …, *p*_l_ with *p*_l_ = the number of miRNA in Reduced signature set

and *y* = 1,2,3, …, *p*_2_ where *p*_2_ = the number of miRNA in Background

and *i* = 1,2,3, …, *n* where *n* = the sample size after filtering

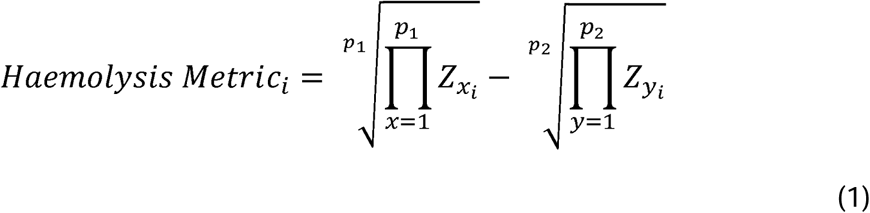

## Application

By using the Haemolysis metric calculation, researchers/clinicians can assess their samples for evidence of haemolysis and obtain recommendations for their own individual samples as clear for use (‘Clear’) or use with caution (‘Caution’). We have shown that when RBC-associated miRNA transcripts are retained in a plasma sample post centrifugation, with subsequent incorporation into the sequencing library, the relative abundance of these miRNAs is increased. This increase is evidenced by the increase in the geometric mean of signature haemolysis miRNAs away from that of the background miRNA giving the background miRNA a smaller relative expression than would be expected from a pure plasma sample taken from the same individual. Using miRNA sequencing data, DraculR is designed to analyse and visualise the distribution of miRNA counts from our haemolysis signature set and compare this with the distribution of counts from background miRNAs. Once calculated, the sample specific Haemolysis metric for user defined data is returned in tabular and graphical format for download and assessment. Samples with a Haemolysis metric ≥ 1.9, our recommended threshold, indicate haemolysis as evidenced by the RBC-associated miRNA retention. Haemolysed samples are identified by the word ‘Caution’ in the ‘Haemolysis Result’ column of the ‘Results Summary’ tab. We recommend removal, or at a minimum further investigation, of any samples that return a Haemolysis metric above the threshold set here prior to use in any downstream analysis.

## Public data example

To illustrate the utility of the application we downloaded four publicly available human plasma HTS miRNA datasets from NCBI GEO (21). The datasets used here were GSE153813, GSE118038, GSE105052, GSE151341 (22–24). Where the publication associated with the given dataset included miRNA differentially expressed between the conditions being considered, and these miRNA correspond with our haemolysis signature set, these miRNA were dropped from the Haemolysis metric calculation. In each example, we were able to detect evidence of haemolysis in multiple samples (Table 1).

**Table 1.** Publicly available human plasma miRNA data were assessed for haemolysis using the DraculR method identifying multiple samples to use with caution in each dataset. No haemolysis information was included with the original dataset.

| Dataset | Experimental Context | Total Samples | Caution | Differentially abundant miRNA | Publication |
| --- | --- | --- | --- | --- | --- |
| GSE153813 | Case:Control<br>Profile miRNA expression at each stage of menstrual cycle; endometriosis | 9 | 3 | NA | NA |
| GSE118038 | Case:Control<br>Prostate cancer biomarker | 70 | 32 | miR-4732-3p,<br>let-7a,<br>miR-26b-5p,<br>miR-98-5p,<br>miR-30c-5p*<br>miR-21-5p | (24) |
| GSE105052 | Case:Control<br>Friedreich's ataxia | 42 | 3 | miR-128-3p,<br>miR-625-3p,<br>miR-130b-5p,<br>miR-151a-5p,<br>miR-330-3p,<br>miR-323a-3p,<br>miR-142-3p | (22) |
| GSE151341 | Case:Control<br>Early radiographic knee osteoarthritis biomarker | 91 | 4 | miR-335-3p,<br>miR-199a-5p,<br>miR-671-3p,<br>miR-1260b,<br>miR-191-3p,<br>miR-335-5p,<br>miR-543 | (23) |

\* miRNA associated with Haemolysis Metric signature

DraculR provides a visual representation of the results in the form of a histogram (Figure 1). In this histogram, the difference between the geometric means can be seen against a background of data with known haemolysis quantification based on the ΔCq (miR-23a-miR-451) method. Samples are coloured to indicate either ‘Clear’ (light blue) or ‘Caution’ (scarlet) with background data coloured to indicate the ΔCq (miR-23a-miR-451) result of either ‘Clear (ΔCq)’ (grey with a light blue highlight) for samples with ΔCq < 7 or ‘Haemolysed (ΔCq)’ (grey with a scarlet highlight) for samples with ΔCq ≥ 7. Background data is taken from Smith *et al*. (20) and represents individuals for which we have both HTS and ΔCq (miR-23a-miR-451) analyses. The false positives reported here (ie where DraculR reports haemolysis but ΔCq < 7) may not be accurate as ΔCq values may be affected by pregnancy status.

**Figure 1.**
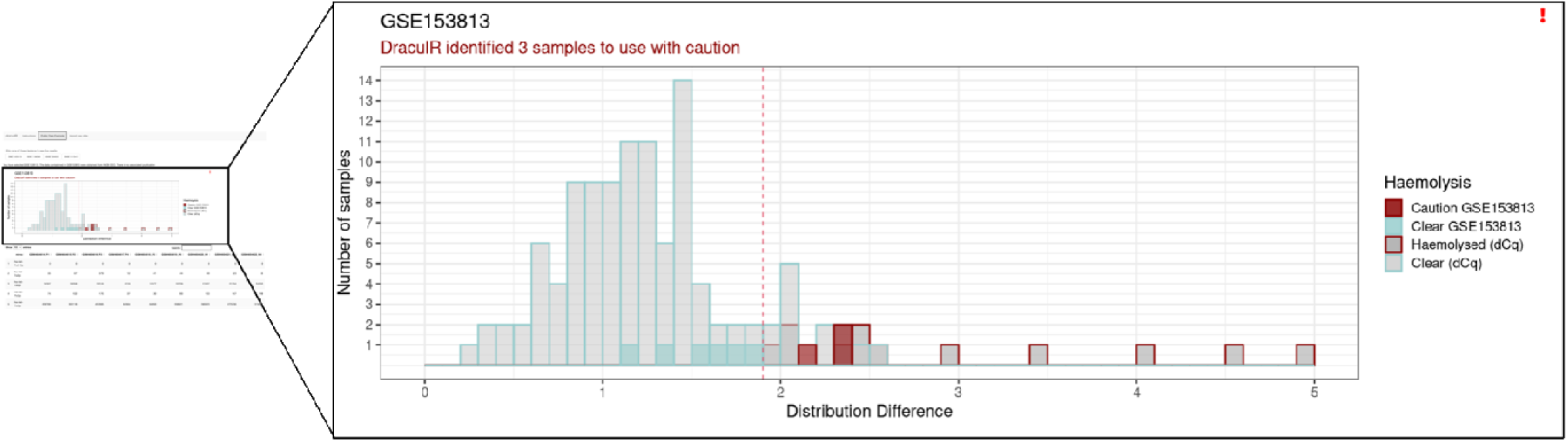
DraculR uses public data to illustrate the potential for unidentified haemolysis with potential to confound biomarker analysis. Here, data pertaining to GSE105052 was retrieved from NCBI Geo for analysis. The screen shot above shows an example where five samples were identified to be used with caution. All data are presented against a background of haemolysed samples assessed using the ΔCq method.

## Conclusion

We have developed a Shiny/R web-based application that allows users to detect and address the issue of haemolysis in plasma miRNA HTS data. DraculR addresses the need for quality control where, either through use of public data, exhaustion of sample, or exhaustion of funds, it is not possible to assess haemolysis using one of the current gold standard approaches (being delta quantification cycle (Cq) values for miR-23a-miR-451 or Spectrophotometry for haemoglobin estimation). The application is easy to use and applicable to small RNA sequencing data from human plasma. The method is robust for cases where a case-control style analysis is undertaken, and the user has *a priori* knowledge of miRNA that are anticipated to be differentially abundant between groups. Whilst a probabilistic quantification of contamination risk is not possible based on the dataset used here, we plan future work drawing on the methods used by Shah *et al*. (2016) that will include serial dilution and miRNA quantification of haemolysed plasma samples to validate and further refine our method. DraculR adds value to the growing resource of public data shared by plasma researchers by enabling *in silico* analysis of haemolysis confounding post sequencing. The detection of haemolysis using our Haemolysis metric enables the user to identify and potentially discard low quality samples which are otherwise unknown to be affected by haemolysis. This enables an additional quality metric and the subsequent increased confidence in the use of high throughput miRNA sequencing data for which no haemolysis information is available (for more details see Supplementary Material).

## Data Availability

All data produced in the present study are available upon reasonable request to the authors or will be released on publication of the primary research article associated with this software note.

## Acknowledgements

We wish to acknowledge the generosity of the women who donated their blood for our research. Without them, this research would not be possible. We also acknowledge valuable input from QIAGEN Genomic Services.

## Funding information

This research is supported by NIH NICHD R01 [grant number HD089685-01] Maternal molecular profiles reflect placental function and development across gestation PI Roberts, an Australian Government Research Training Program (RTP) Scholarship awarded to MDS, a National Health and Medical Research Council Investigator Grant [grant number GNT1174971] awarded to CTR and a Matthew Flinders Professorial Fellowship awarded to CTR and funded by Flinders University. JB is supported by the James & Diana Ramsay Foundation. KAP is supported by the Florey Fellowship funded by the Adelaide Hospital Research Committee.

## Notes

### Competing Interest Statement

The authors have declared no competing interest.

### Author Declarations

Ethics approval for the collection of blood from women undergoing elective pregnancy termination between 6 to 23 weeks of gestation was provided under HREC 16 TQEH 33, by The Queen Elizabeth Hospital Human Research Ethics Committee (TQEH LMH MH). Blood from women forming the general population group was collected after informed consent with ethics approval provided under HREC H 021 2005, by The University of Adelaide Human Research Ethics Committee.

## References

1. Pritchard CC, Cheng HH, Tewari M. MicroRNA profiling: Approaches and considerations [Internet]. Vol. 13, Nature Reviews Genetics. NIH Public Access; 2012. p. 358–69. Available from: http://www.pubmedcentral.nih.gov/articlerender.fcgi?artid=PMC4517822

2. Guo R, Fan G, Zhang J, Wu C, Du Y, Ye H, et al. A 9-microRNA Signature in Serum Serves as a Noninvasive Biomarker in Early Diagnosis of Alzheimer’s Disease. J Alzheimers Dis. 2017;60(4):1365–77.

3. Sánchez-Mora C, Soler Artigas M, Garcia-Martínez I, Pagerols M, Rovira P, Richarte V, et al. Epigenetic signature for attention-deficit/hyperactivity disorder: identification of miR-26b-5p, miR-185-5p, and miR-191-5p as potential biomarkers in peripheral blood mononuclear cells. Neuropsychopharmacology. 2019 Apr;44(5):890–7.

4. Jin X, Chen Y, Chen H, Fei S, Chen D, Cai X, et al. Evaluation of Tumor-Derived Exosomal miRNA as Potential Diagnostic Biomarkers for Early-Stage Non-Small Cell Lung Cancer Using Next-Generation Sequencing. Clin Cancer Res. 2017 Sep 1;23(17):5311–9.

5. Coenen-Stass AML, Magen I, Brooks T, Ben-Dov IZ, Greensmith L, Hornstein E, et al. Evaluation of methodologies for microRNA biomarker detection by next generation sequencing. RNA Biol. 2018 Sep 18;15(8):1133–45.

6. Williams Z, Ben-Dov IZ, Elias R, Mihailovic A, Brown M, Rosenwaks Z, et al. Comprehensive profiling of circulating microRNA via small RNA sequencing of cDNA libraries reveals biomarker potential and limitations. Proceedings of the National Academy of Sciences [Internet]. 2013; Available from: http://dx.doi.org/10.1073/pnas.1214046110

7. Keller A, Leidinger P, Steinmeyer F, Stähler C, Franke A, Hemmrich-Stanisak G, et al. Comprehensive analysis of microRNA profiles in multiple sclerosis including next-generation sequencing. Mult Scler. 2014 Mar;20(3):295–303.

8. Saini J, Bandyopadhyay B, Pandey AD, Ramachandran VG, Das S, Sood V, et al. High-Throughput RNA Sequencing Analysis of Plasma Samples Reveals Circulating microRNA Signatures with Biomarker Potential in Dengue Disease Progression. mSystems [Internet]. 2020 Sep 15;5(5). Available from: http://dx.doi.org/10.1128/mSystems.00724-20

9. Yu F, Pillman KA, Neilsen CT, Toubia J, Lawrence DM, Tsykin A, et al. Naturally existing isoforms of miR-222 have distinct functions. Nucleic Acids Res. 2017 Nov 2;45(19):11371–85.

10. Pillman KA, Goodall GJ, Bracken CP, Gantier MP. miRNA length variation during macrophage stimulation confounds the interpretation of results: implications for miRNA quantification by RT-qPCR. RNA. 2019 Feb;25(2):232–8.

11. Smith MD, Pillman K, Jankovic-Karasoulos T, McAninch D, Wan Q, Bogias KJ, et al. Large-scale transcriptome-wide profiling of microRNAs in human placenta and maternal plasma at early to mid gestation. RNA Biol. 2021 Aug 19;1–14.

12. Kirschner MB, Kao SC, Edelman JJ, Armstrong NJ, Vallely MP, van Zandwijk N, et al. Haemolysis during sample preparation alters microRNA content of plasma. Pfeffer S, editor. PLoS One. 2011 Sep 1;6(9):e24145.

13. Pritchard CC, Kroh E, Wood B, Arroyo JD, Dougherty KJ, Miyaji MM, et al. Blood cell origin of circulating microRNAs: a cautionary note for cancer biomarker studies. Cancer Prev Res. 2012 Mar;5(3):492–7.

14. Blondal T, Jensby Nielsen S, Baker A, Andreasen D, Mouritzen P, Wrang Teilum M, et al. Assessing sample and miRNA profile quality in serum and plasma or other biofluids [Internet]. Vol. 59, Methods. Academic Press; 2013. p. 164–9. Available from: https://www.sciencedirect.com/science/article/pii/S1046202312002551

15. Kirschner MB, Edelman JJB, Kao SCH, Vallely MP, Van Zandwijk N, Reid G. The impact of hemolysis on cell-free microRNA biomarkers. Front Genet. 2013;4(MAY):94.

16. Sun L, Yu Y, Niu B, Wang D. Red Blood Cells as Potential Repositories of MicroRNAs in the Circulatory System. Front Genet. 2020 Jun 3;11:442.

17. Cheng HH, Yi HS, Kim Y, Kroh EM, Chien JW, Eaton KD, et al. Plasma Processing Conditions Substantially Influence Circulating microRNA Biomarker Levels. Kiechl S, editor. PLoS One. 2013 Jun 7;8(6):e64795.

18. Livak KJ, Schmittgen TD. Analysis of Relative Gene Expression Data Using Real-Time Quantitative PCR and the 2-ΔΔCT Method [Internet]. Vol. 25, Methods. 2001. p. 402–8. Available from: http://dx.doi.org/10.1006/meth.2001.1262

19. Wong C-H, Song C, Heng K-S, Kee IHC, Tien S-L, Kumarasinghe P, et al. Plasma free hemoglobin: a novel diagnostic test for assessment of the depth of burn injury. Plast Reconstr Surg. 2006 Apr;117(4):1206–13.

20. Melanie D. Smith, Shalem Y. Leemaqz, Tanja Jankovic-Karasoulos, Dale McAninch, Dylan McCullough, James Breen, Claire T. Roberts, Katherine A. Pillman. Haemolysis detection in microRNA-seq from clinical plasma samples. MedRxiv.

21. Edgar R, Domrachev M, Lash AE. Gene Expression Omnibus: NCBI gene expression and hybridization array data repository. Nucleic Acids Res. 2002 Jan 1;30(1):207–10.

22. Seco-Cervera M, González-Rodríguez D, Ibáñez-Cabellos JS, Peiró-Chova L, Pallardó FV, García-Giménez JL. Small RNA-seq analysis of circulating miRNAs to identify phenotypic variability in Friedreich’s ataxia patients. Sci Data. 2018 Mar 6;5:180021.

23. Ali SA, Gandhi R, Potla P, Keshavarzi S, Espin-Garcia O, Shestopaloff K, et al. Sequencing identifies a distinct signature of circulating microRNAs in early radiographic knee osteoarthritis. Osteoarthritis Cartilage. 2020 Nov;28(11):1471–81.

24. Giglio S, De Nunzio C, Cirombella R, Stoppacciaro A, Faruq O, Volinia S, et al. A preliminary study of micro-RNAs as minimally invasive biomarkers for the diagnosis of prostate cancer patients. J Exp Clin Cancer Res. 2021 Feb 23;40(1):79.

