## Supplemental File 1 for "DraculR: A web based application for *in silico* haemolysis detection in high throughput small RNA sequencing data"

### Supplementary Data

The Haemolysis metric described here (1) and implemented in the DraculR Shiny/R web-based application, performs an *in silico* quality assessment to detect evidence of haemolysis contamination in the original plasma specimen, assigning each small RNA sequencing dataset into one of two categories. The classification of ‘Clear’ or ‘Caution’ alert the user to potential quality control issues in specimen.

#### Application

DraculR provides a simple GUI interface that allows the user to upload small RNA sequencing data in the form of a raw counts table with names in mature miRNA format, i.e. miR-106b-3p (2) (Figure 1a). The interface provides the option to personalise table and figure titles (Figure 1b), set filtering options (Figure 1c) and remove miRNA known to be differentially abundant between the user groups of interest from the calculation of the Haemolysis metric (Figure 1d). This final step ensures the calculation for haemolysis is not confounded in the event that one or more of the miRNA signature set is known to differ between groups.

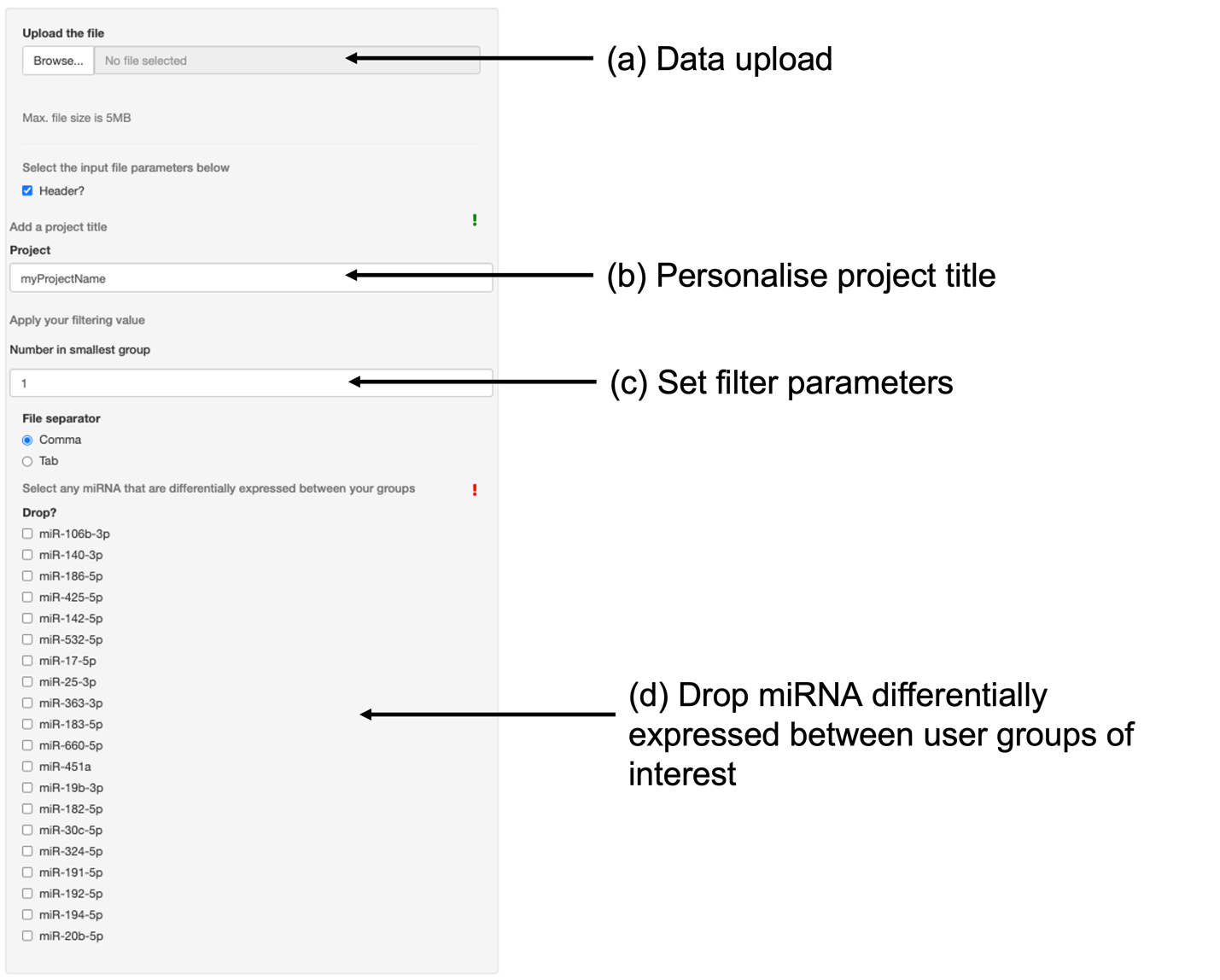

**Figure 1**. DraculR provides a simple GUI interface for data upload and manipulation. First, the user selects a local file for upload (a) and changes the project title to be used on tables and figures (b). Prior to the algorithm running, the user then selects the minimum number of samples per group for filtering (c) and selects any miRNA which are known to be differentially abundant between the user groups of interest (d) to be dropped from the Haemolysis metric calculation.

In Figure 2 we present an example distribution illustrative of a dataset classified as Clear (Figure 2a) and as one as Caution (Figure 2b). In the first example (Figure 2a), the distance between the geometric mean of the Background miRNA (light blue) compared to that of the Classifier miRNA (scarlet) is very small suggesting the two sets of miRNA belong to the same distribution. In the second example (Figure 2b) the distance between the geometric mean of the Background miRNA compared to that of the Classifier miRNA is larger than the ‘Clear’ example. Furthermore, this difference is greater than the Haemolysis Metric threshold of 1.9, established in Smith *et al.* (1), and thus the sample has been classified as ‘Caution’. In Figure 2b the Background and Classifier distributions appear to be independent. This suggests that RBC-associated miRNA have been added to the pool of miRNA isolated in the plasma. In the haemolysed example (Figure 2b), we would recommend removing the sample data from further analysis. However, where a decision is made to retain samples, the issue of haemolysis should be noted and may be a limitation to inference.

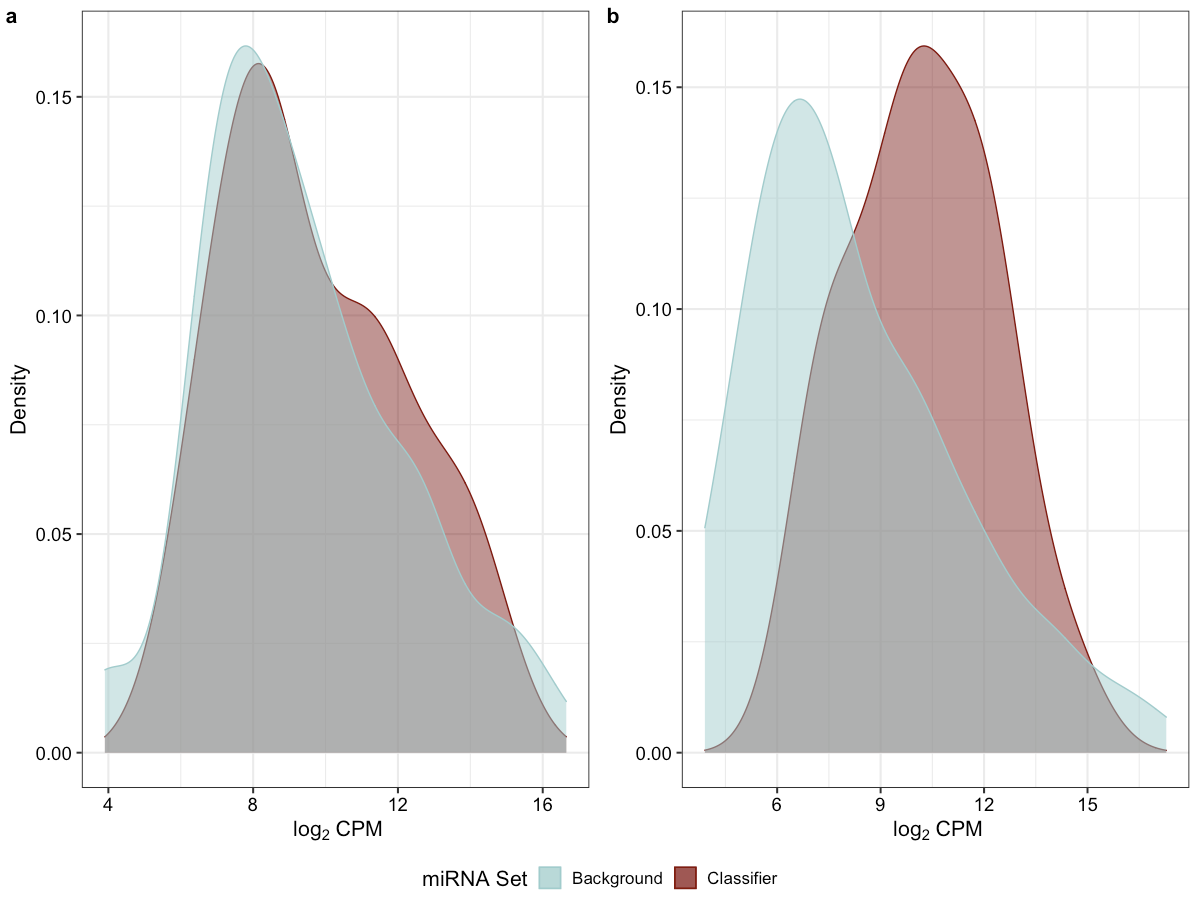

**Figure 2**. In the first example (a) the sample is classified as ‘Clear’ indicating no evidence for haemolysis. The distance between the geometric mean of Background and Classifier miRNA is small. In the second example (b) the sample is classified as ‘Caution’ indicating that we found evidence suggestive of haemolysis. The geometric mean of Background and Classifier miRNA is further apart than that expected where no haemolysis is present, with the Haemolysis Metric ≥ than the threshold of 1.9.

#### Inputting data into DraculR

DraculR allows the user to upload a raw, high throughput sequencing counts table for analysis. Whilst normalisation is locked to the Trimmed Mean of M method (TMM) previously recommended in the *edgeR* workflow (3), the user controls features such as filtering for low expression (Figure 3) and refining the haemolysis signature set based on *a priori* knowledge of miRNAs that may be differentially expressed in the comparison of interest (Figure 4). The purpose of removing miRNAs with a known association to the condition of interest is to help ensure any issues with haemolysis are not confounded with the research hypothesis. Note that samples with total miRNA read counts < 1 million are considered to be poorly sequenced and are recommended to be removed for quality control.

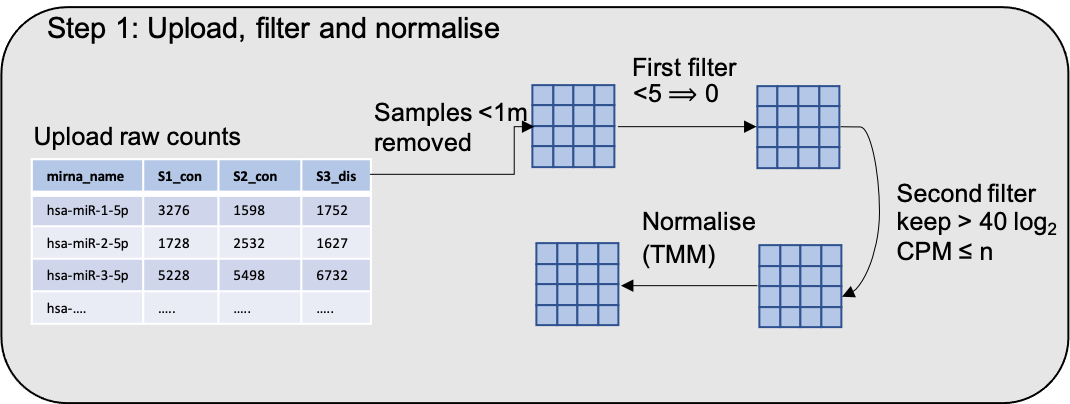

**Figure 3**. Import a raw counts table generated by high throughput miRNA sequencing of human plasma libraries. These data will be filtered according to user specified requirements (n = number of samples in the smallest group of interest) and normalised using the Trimmed Mean of M (TMM) method as previously recommended in the *edgeR* workflow (3).

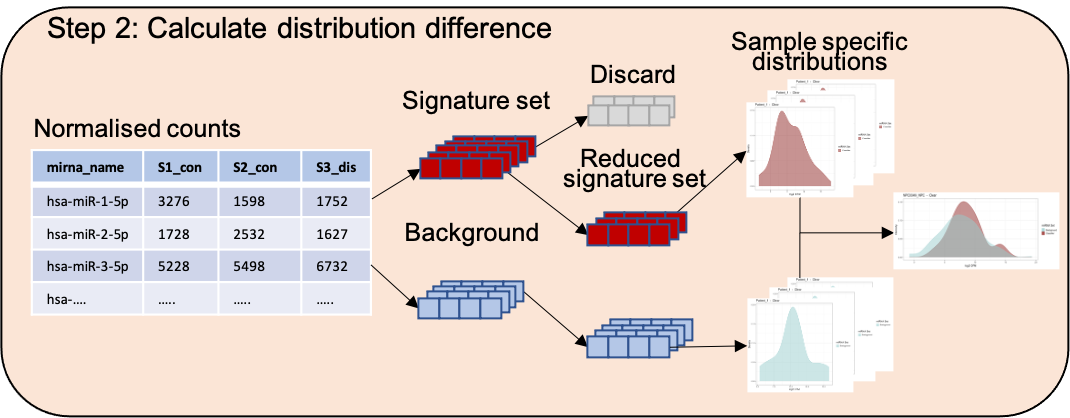

**Figure 4**. The distribution difference between the background and signature miRNA counts is calculated on an individual sample basis allowing the user to upload one to many samples as required. In the case of *a priori* knowledge of miRNA differentially abundant between a tested condition/control paradigm the user may choose to reduce the signature miRNA such that they do not include miRNA of interest (recommended).

#### Visualisation and interpretation

An essential feature of DraculR is that it allows users to visualise and assess the values obtained in the results, through sample specific and consolidated graphics including density plots, histograms and tables (Figure 5). These features help the user decide on the level of haemolysis that may affect their analysis by providing a new quality metric. Using this metric the user may choose to remove samples from downstream analyses. However, irrespective of whether samples with a Haemolysis metric above the suggested threshold are removed or retained, the new information may be important to the analysis of their miRNA sequencing data.

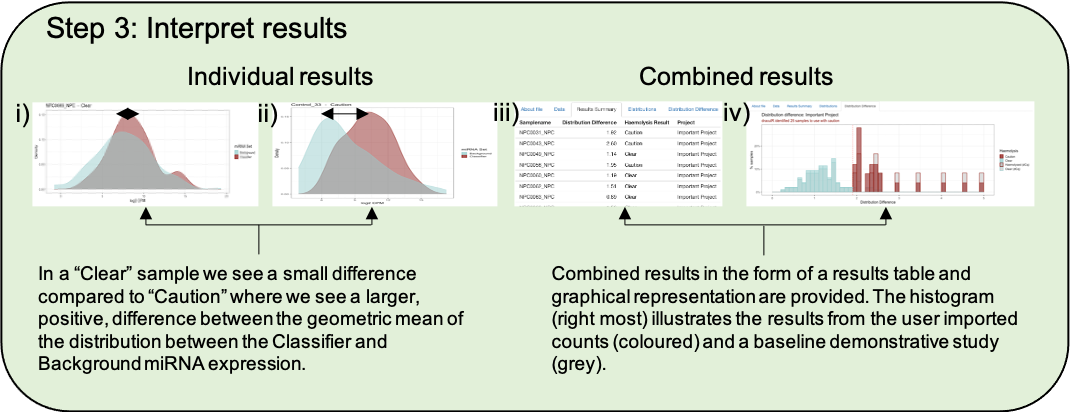

**Figure 5**. Graphical results in the form of a density plot of individual distributions (i, ii) and a histogram of combined distribution differences (iv) are provided along with a combined table of results (iii). The user is provided with both a metric describing the amount of haemolysis and a recommendation of caution, if appropriate (iii).

#### Public data example

To illustrate the utility of the application, we downloaded four publicly available human plasma high throughput sequencing miRNA datasets from NCBI GEO (4). The datasets used here were GSE153813, GSE118038, GSE105052, GSE151341 (5–7). Each of the publicly available datasets was successfully processed through DraculR.

In GSE153813, RNA libraries were prepared using human plasma and expression profiles were generated using an Illumina NextSeq 500. The aim of the study was to profile miRNA expression across the menstrual cycle in the context of endometriosis, a disorder characterised by the growth of uterine tissue outside of the uterus. Given the nature of the experiment, all participants were female. Samples were grouped by health status, Endometriosis (n = 6) and Healthy (n = 3). An average of ~2.2 million reads (Figure 6) were sequenced per sample (range 955209 to 3815839 reads). All libraries were retained for analysis using DraculR, which identified 3 samples to be used with caution in downstream analysis (Figure 7).

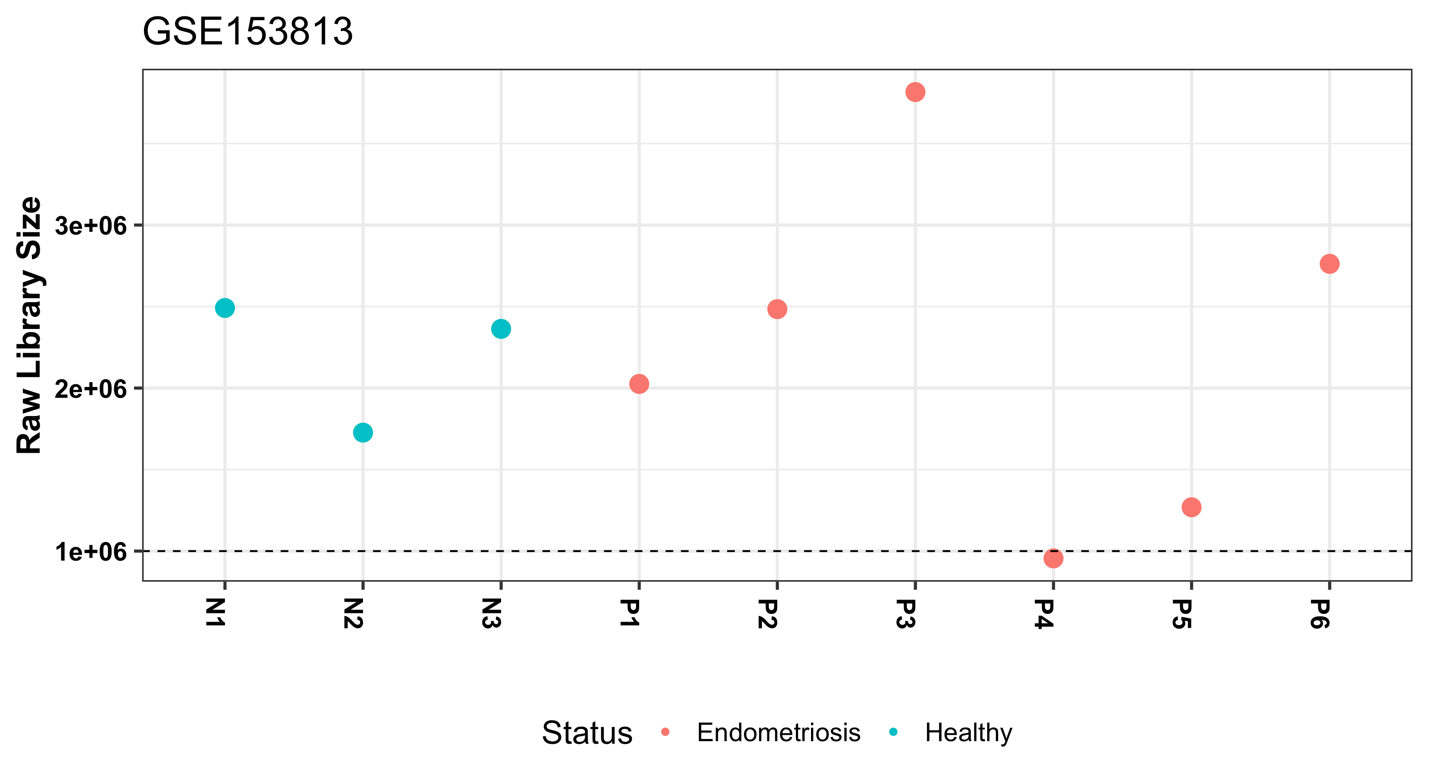

**Figure 6**. Sequencing read depth for GSE153813. Dashed line represents one million reads.

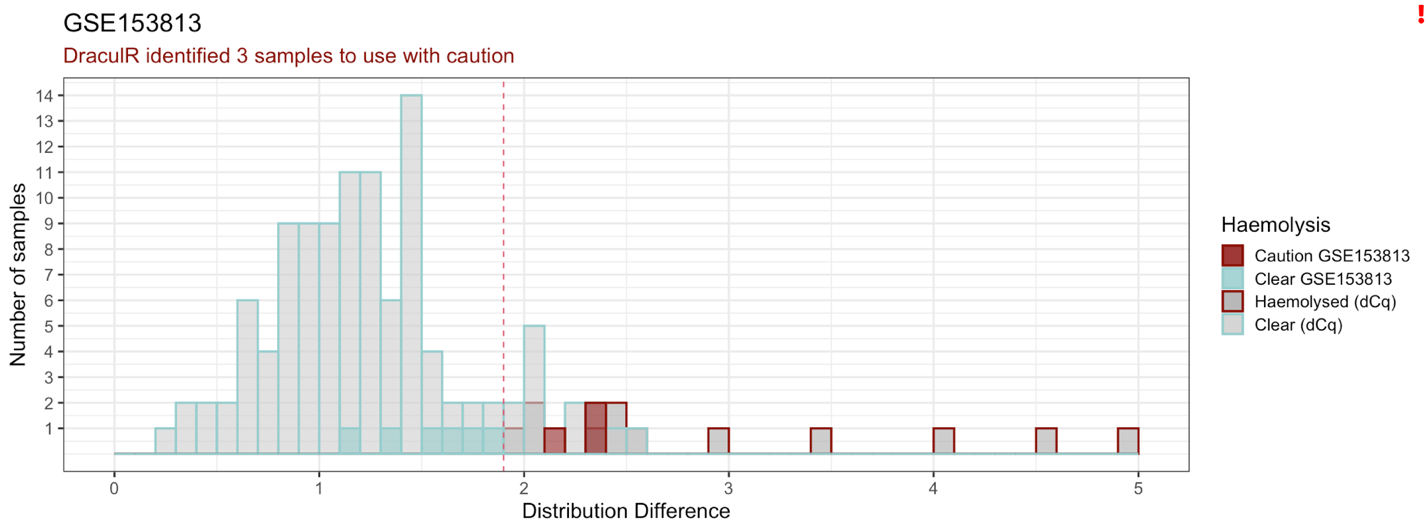

**Figure 7**. DraculR identified 3 samples from GSE153813 to be used with caution in downstream analysis.

In GSE118038, RNA libraries were prepared using human plasma and expression profiles were generated using an Illumina NextSeq 500. The aim of this study was to explore the biomarker potential of plasma derived miRNAs in the context of prostate cancer diagnosis (7). Given the nature of the experiment, all participants were male. Samples were grouped by health status, Prostate Cancer (n = 33) and Healthy (n = 37). An average of ~0.7 million reads (Figure 8) were sequenced per sample (range 207,744 to 1,915,974 reads). This study identified six miRNA that were differentially abundant between healthy individuals and those diagnosed with prostate cancer (Table 1 of the main paper). Of these, one, miR-30c-5p, forms part of the 20 miRNA signature set and was dropped from the Haemolysis metric calculation using the option shown in Figure 1b. All libraries were retained for analysis using DraculR, which identified 32 samples to be used with caution in downstream analysis (Figure 9).

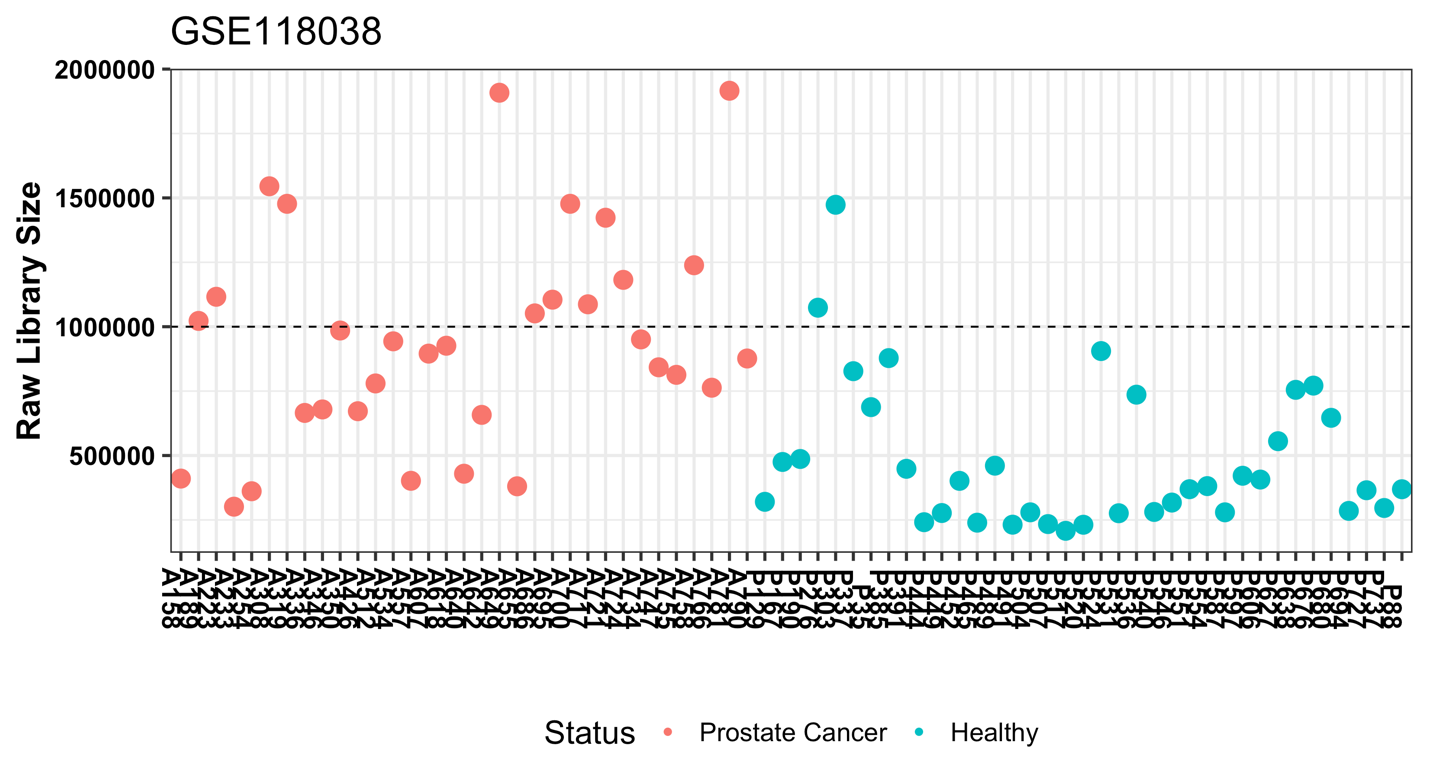

**Figure 8**. Sequencing read depth for GSE118038. Dashed line represents one million reads.

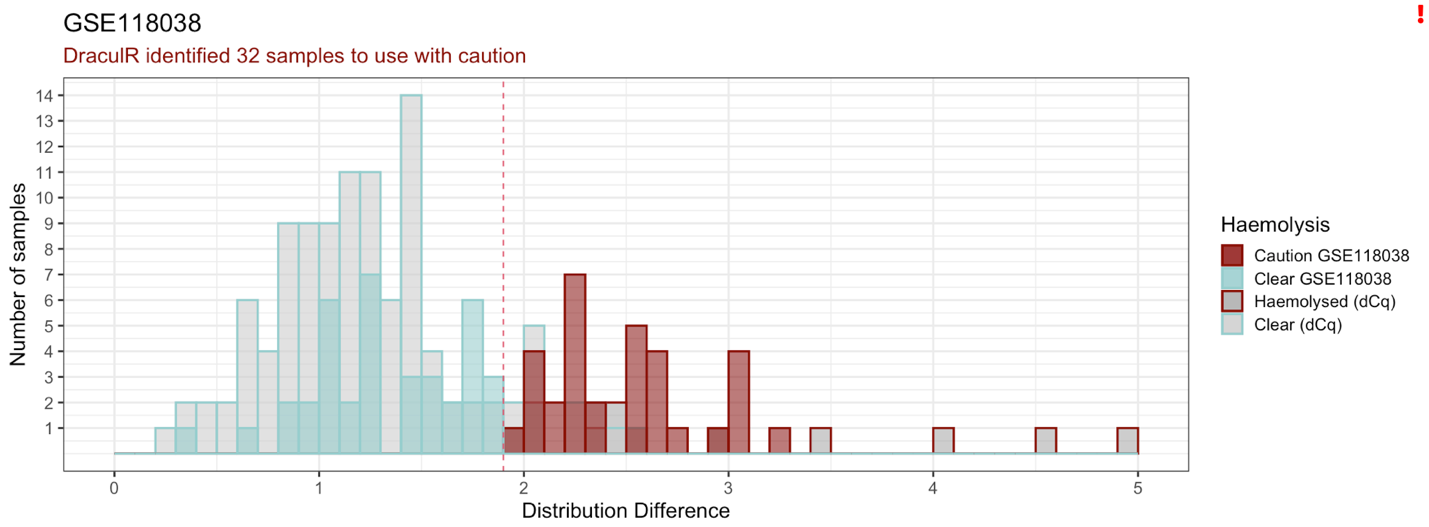

**Figure 9**. DraculR identified 32 samples from GSE118038 to be used with caution in downstream analysis.

In GSE105052, RNA libraries were prepared using human plasma and expression profiles were generated using an Illumina HiScanSQ. The aim of this study was to explore the biomarker potential of plasma derived miRNAs in the context of Friedreich’s ataxia (5). Samples were grouped by health status, Friedreich’s ataxia (n = 25) and Healthy (n = 17). An average of ~0.13 million reads (Figure 10) were sequenced per sample (range 1,474 to 695,556 reads). This study identified seven miRNA that were differentially abundant between healthy individuals and those diagnosed with Friedreich’s ataxia (Table 1 of the main paper), none of which is part of the 20 miRNA signature set used in the Haemolysis metric calculation. All libraries were retained for analysis using DraculR which identified three samples to be used with caution in downstream analysis (Figure 11).

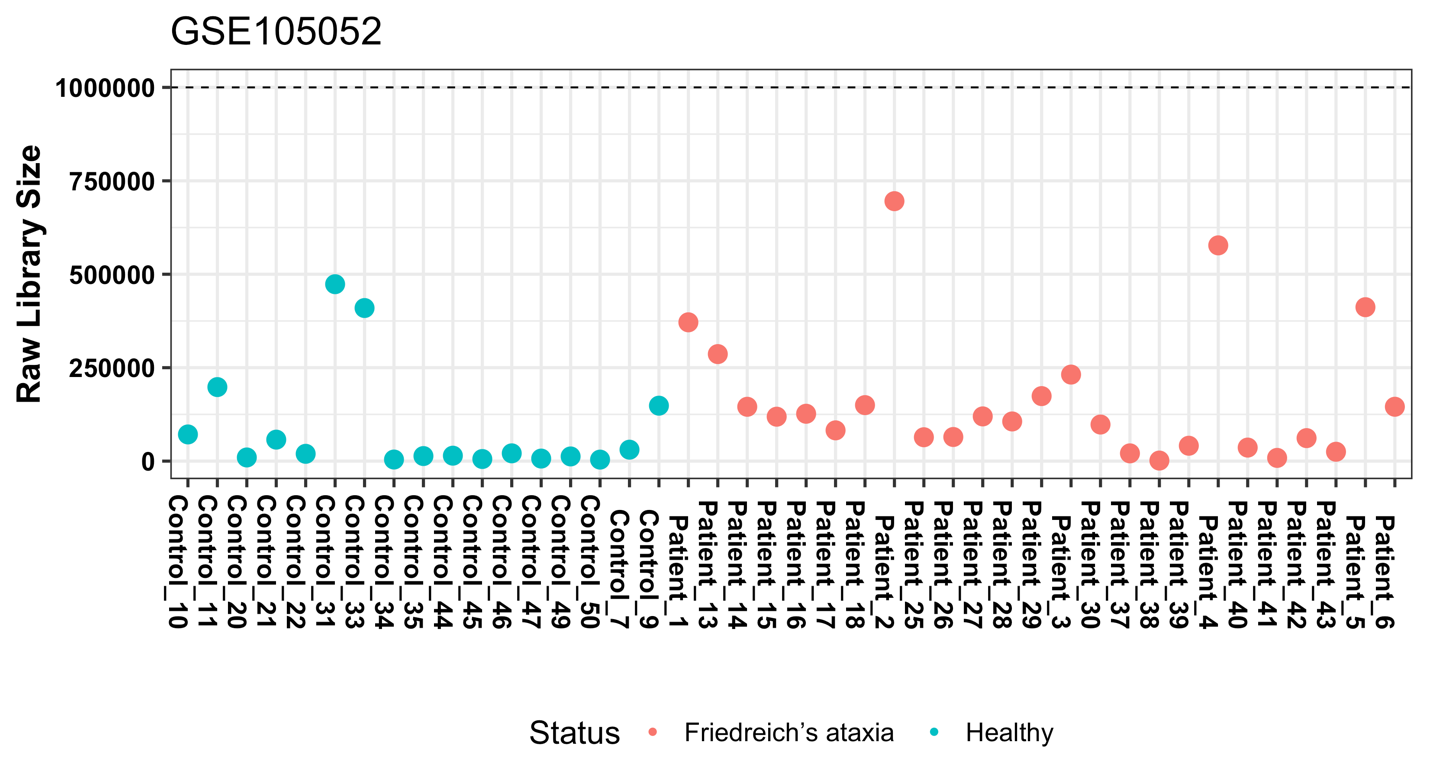

**Figure 10**. Sequencing read depth for GSE105052. Dashed line represents one million reads.

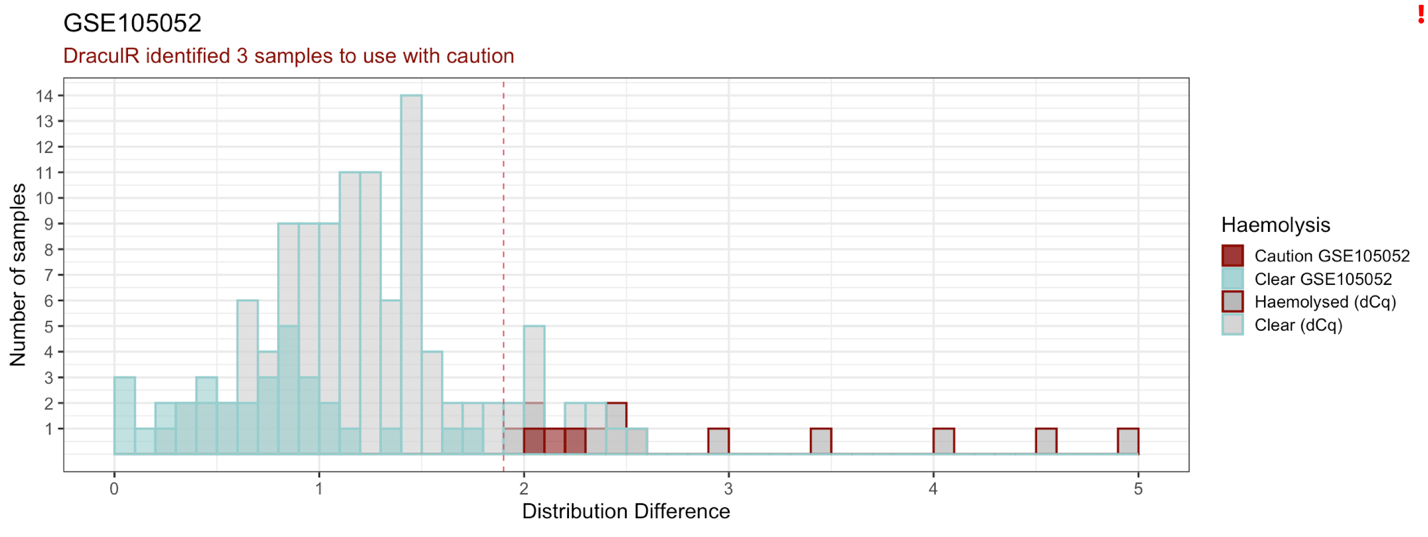

**Figure 11**. DraculR identified 3 samples from GSE105052 to be used with caution in downstream analysis.

In GSE151341, RNA libraries were prepared using human plasma and expression profiles were generated using an Illumina NextSeq 550 (6). The aim of the study was to sequence plasma miRNAs from patients with radiographic knee osteoarthritis to identify unique miRNA signatures in each of two disease states. Samples were grouped by disease status, early [Kellgren-Lawrence grade 0 or 1 (n=41)] or late [Kellgren-Lawrence grade 3 or 4 (n=50)] symptomatic radiographic knee osteoarthritis. An average of ~2 million reads (Figure 12) was sequenced per sample (range 396,722 to 6,092,971 reads). This study identified seven miRNA that were differentially abundant between individuals diagnosed with early or late radiographic knee osteoarthritis (Table 1 of the main paper), none of which was part of the 20 miRNA signature set used in the Haemolysis metric calculation. All libraries were retained for analysis using DraculR which identified three samples to be used with caution in downstream analysis (Figure 13).

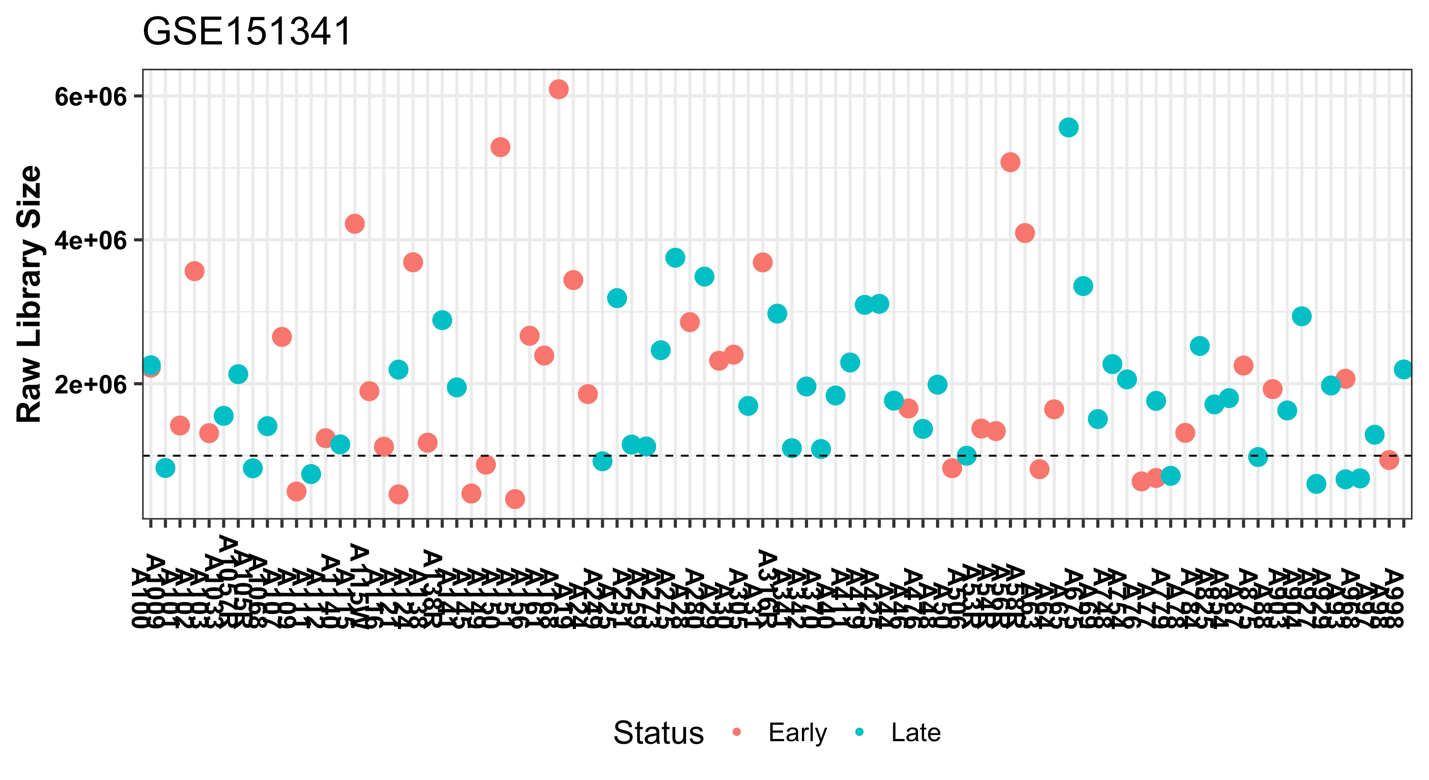

**Figure 12**. Sequencing read depth for GSE151341. Dashed line represents one million reads.

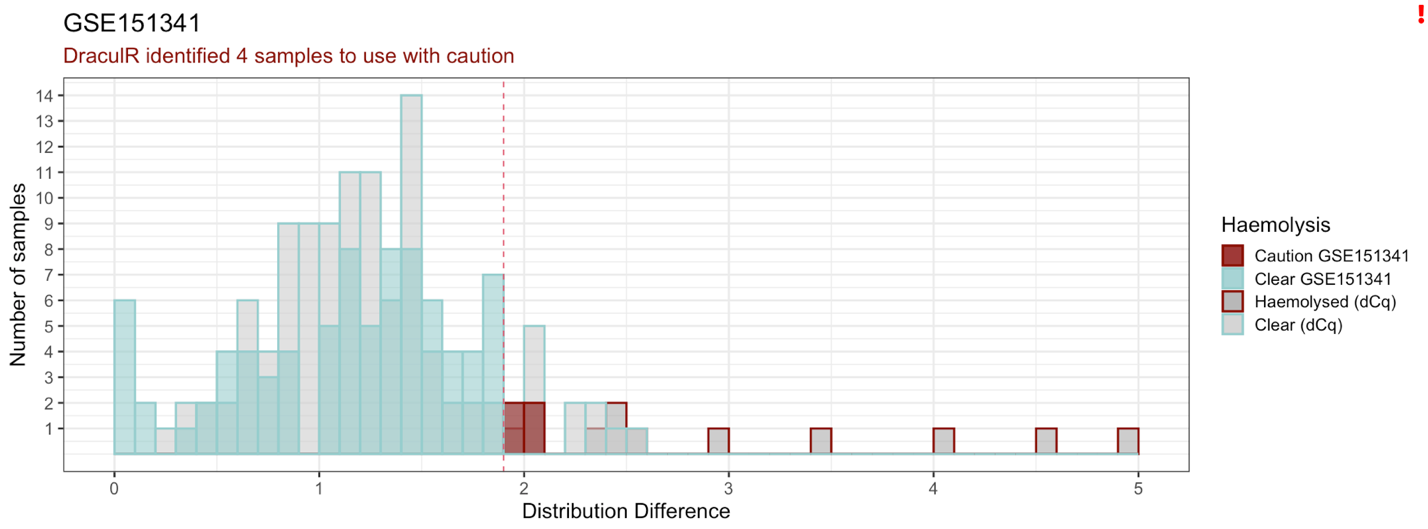

**Figure 13**. DraculR identified 4 samples from GSE105052 to be used with caution in downstream analysis.

Detailed information regarding each of the public data sets can be found in Table 1 of the main paper.

References

1.	Melanie D. Smith, Shalem Y. Leemaqz, Tanja Jankovic-Karasoulos, Dale McAninch, Dylan McCullough, James Breen, Claire T. Roberts, Katherine A. Pillman. Haemolysis detection in microRNA-seq from clinical plasma samples. MedRxiv.

2.	Kozomara A, Griffiths-Jones S. miRBase: integrating microRNA annotation and deep-sequencing data. Nucleic Acids Res. 2011 Jan;39(Database issue):D152-7.

3.	Robinson MD, McCarthy DJ, Smyth GK. edgeR: A Bioconductor package for differential expression analysis of digital gene expression data. Bioinformatics. 2009;26(1):139–40.

4.	Edgar R, Domrachev M, Lash AE. Gene Expression Omnibus: NCBI gene expression and hybridization array data repository. Nucleic Acids Res. 2002 Jan 1;30(1):207–10.

5.	Seco-Cervera M, González-Rodríguez D, Ibáñez-Cabellos JS, Peiró-Chova L, Pallardó FV, García-Giménez JL. Small RNA-seq analysis of circulating miRNAs to identify phenotypic variability in Friedreich’s ataxia patients. Sci Data. 2018 Mar 6;5:180021.

6.	Ali SA, Gandhi R, Potla P, Keshavarzi S, Espin-Garcia O, Shestopaloff K, et al. Sequencing identifies a distinct signature of circulating microRNAs in early radiographic knee osteoarthritis. Osteoarthritis Cartilage. 2020 Nov;28(11):1471–81.

7.	Giglio S, De Nunzio C, Cirombella R, Stoppacciaro A, Faruq O, Volinia S, et al. A preliminary study of micro-RNAs as minimally invasive biomarkers for the diagnosis of prostate cancer patients. J Exp Clin Cancer Res. 2021 Feb 23;40(1):79.
